# The integrated approach of learning tuberculosis transmission within and outside households via random directed graph models

**DOI:** 10.1101/2020.07.30.20165506

**Authors:** Tenglong Li, Edward C. Jones-L□pez, Laura F. White

## Abstract

Household contact studies are frequently used in tuberculosis transmission research, and models based on them often focus on transmission within the household. This contradicts recent research which suggests the transmission may be more likely to happen outside the household than within the household in high burden settings where these studies are frequently conducted. Consequently, most models would lead to biased estimates and misleading public health interventions. There is a strong need for developing models that allow concurrent estimation of household and extra-household transmission. In this study, we develop a random directed graph model for tuberculosis transmission, which permits users to concurrently build models for both household and extra-household transmission. Furthermore, our model can estimate the relative frequency of household transmission versus extra-household transmission and consistently produce unbiased estimates for risk factors, regardless of whether community controls are available. We illustrate our approach with a household contact study conducted in Vitória, Brazil, and our results indicate that extra-household transmission can account for 63% to 98% of *M. tuberculosis* infections detected during such a study.

## 1 Introduction

Tuberculosis (TB) is one of the oldest infectious diseases, yet it remains a major threat to global health.^1,2^ One obstacle for global TB control is limited knowledge about TB transmission dynamics, which is complicated by long latency, imperfect TB diagnostics as well as the uncertainty about the determinants of progression to active TB disease.^3,4^ Household contact studies, which enroll newly diagnosed pulmonary TB patients (index cases) and their household contacts, are often used to study TB transmission.^5-7^ Inferences based on household contact studies seldom consider extra-household transmission,^8,9^ which contradicts the findings of a growing body of literature that suggests extra-household transmission is even more substantial than household transmission in high burden settings.^10,11^ Consequently, such inferences may bias estimates of the general transmission dynamics of TB as well as the nature and strength of the associations between risk factors for latent TB infection (LTBI).^12^ For this reason, models that allow concurrent estimation of household and extra-household transmission are important to consider.

Few models exist to concurrently account for household and extra-household transmission of disease. The unified probability model (UPM) has been applied to household contact studies of TB for concurrent estimation of household and extra-household transmission of TB.^4^ Built on a mixed effects modeling structure, it estimates the association between LTBI and exposed host, infector, and environmental risk factors as fixed effects while controlling for household random effects. The UPM additionally has a parameter that characterizes the probability of extra-household transmission so that it can separate the probability of extra-household transmission from the probability of household transmission. Nevertheless, the UPM has three major limitations. First, the extra-household transmission probability may still be non-identifiable due to the fact that it is a part of the intercept in the model, though the non-identifiability issue could be ameliorated by controlling for the most powerful predictors of household transmission. Second, the UPM does not easily model risk factors associated with extra-household transmission. Third, the UPM considers the index cases as the only source of household transmission, which is likely not realistic for households with other active TB diseased cases (called secondary TB diseased cases).

Another approach for modeling household and extra-household sources of infection is the random directed graph model (RDM).^13^ RDMs have proven to be informative in household contact studies for influenza research.^14,15^ Using Markov Chain Monte Carlo (MCMC), RDM concurrently estimates extra-household and household transmission by modeling and imputing the transmission paths outside and within households. RDMs allow one to incorporate factors associated with extra-household transmission and allow all infected cases to be sources of household transmission. This model has typically been implemented in settings where community controls, i.e., households without an index case, are enrolled, creating important data to inform extra-household transmission paths along with serological data.

Household contact studies for TB do not typically enroll community controls and serology data is not relevant for TB. Given community controls play a vital role in distinguishing extra-household transmission from household transmission, RDMs may not be identifiable for household contact studies for TB without modification. Furthermore, RDMs have been developed in the context of influenza which has distinct transmission patterns from TB, so a modified RDM is needed.

We have two aims in this paper. First, we develop an RDM that is modified for TB household contact studies. The central task in this aim is to formalize the rules of drawing random directed graphs for TB household contact studies. The second aim is to explore strategies to solve the non-identifiability issue for RDMs and TB household contact studies. We present three possible strategies: 1) include extremely predictive covariates in the models of extra-household and household transmission; 2) enroll community controls; or 3) use random weights in the MCMC algorithm where the weights serve as prior information for updating transmission paths outside or within a household. We describe how different weighting schemes can be evaluated based on deviance information criterion (DIC).

Our paper is structured as follows: In section 2, we describe our motivating data example, a Brazilian household contact study of TB, as well as the traditional RDM in influenza research. In section 3, we detail a modified RDM for TB household contact studies as well as the weighted MCMC approach for estimation. In section 4, we present the results of two simulation studies as well as the Brazilian household contact study. Section 5 concludes the paper with a discussion of our findings.

## 2 Background

### 2.1 Brazilian household contact study

Led by the “US-Brazil Research Collaboration on Strain Variation in TB” study group, a research initiative under the US National Institute of Health and a part of the International Collaboration in Infectious Diseases Research program, the Brazilian household contact study targeted the population living in the greater metropolitan area of Vitória, Brazil. This study enrolled 160 households, each with an index case who fulfilled the following criteria: 1) age ≥ 18 years with coughing ≥ 3 weeks; 2) a sputum test result of acid-fast bacilli (AFB) ≥ 2+ with subsequent *M. tuberculosis* growth in culture; 3) has no less than three household contacts. This study excluded index TB cases who had been infected with HIV (or refused to be tested for HIV) or treated for TB disease in the past. A household contact in this study was defined to be an individual of any age that had close contact with the index case for more than 3 months. Demographic and environmental information on all 160 index cases and their 934 household contacts has been collected and LTBI status was determined by tuberculin skin test (TST) reading(s) conducted by staff trained by the National TB Program. The household contacts who had TST readings ≥ 10 mm by 8 weeks after enrollment of the index case were considered LTBI according to TST study criteria. Fifty-five household contacts had active TB disease in addition to the initial index cases during the six-years of follow-up visits. The dates of diagnoses for cases who had active TB disease were recorded. In total, the study enrolled 160 households in four municipalities and had 215 cases that had active TB disease and 545 LTBI cases. The study protocol has been described in detail elsewhere.^6,16^

### 2.2 Random directed graph model

Developed in influenza transmission research, the random directed graph model (RDM) considers the unobserved transmission paths as missing data and imputes them using MCMC.^14^ Specifically, for each household one can create a random directed graph where its vertices represent the household members and edges represent the possible transmission paths. The directed graph should be consistent with the observed data and follow rules consistent with disease transmission. These rules are:

1. Infected household members, or those with unknown status, can send and receive edge(s) from the other infected household members and/or the community.
2. Infected household members receive at least one edge.
3. Non-infected household members do not receive or send edges.

In general, the posterior distribution for the RDM is given by:

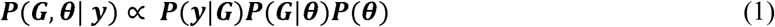

where G denotes the directed graph of the transmission paths for each household, θ represents the transmission model parameters and y is the observed data. P(y|G) equals 1 if the underlying directed graph is consistent with the observed data and 0 otherwise. P(G|θ) is the likelihood of the directed graph given the transmission model and it has the following generic form:

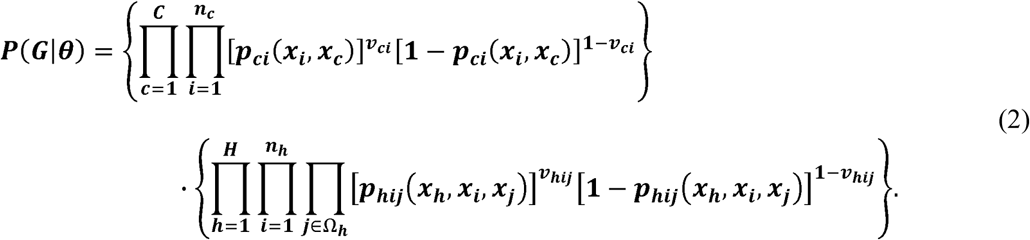

For extra-household transmission, *C* refers to the number of communities enrolled in household contact study and there are *n*_*C*_ participants in the community *c* (*c* = 1,…, *C*). The probability that an infectee *i* is infected by someone outside the household is *p*_*Ci*_(*x*_*i*_, *x*_*C*_), which depends on the features of individual *i, x*_*i*_ and the features of the community of residence, *x*_*C*_. For a directed graph, *ν* _*Ci*_ equals 1 if an edge from the community *c* to the infectee *i* is present or 0 if it is absent. The model for extra-household transmission is

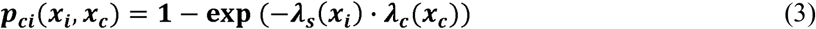

where *λ*_*s*_(*x*_*i*_) and *λ*_*c*_ (*x*_*c*_) characterize the susceptibility of the infectee *i* and the general risk of extra-household transmission in the community *c* respectively.

For household transmission, *H* refers to the number of enrolled households and *n*_*h*_ refers to the number of participants in the household *h* (*h* = 1,…, *H*). The group of infectious participants in the household *h* is denoted as Ω_*h*_. The probability that an infectee *i* is infected by an infector *j* in the household *h* is *p*_*hij*_(*x*_*h*_, *x*_*i*_, *x*_*j*_), which is determined by the features of the household *h*, the infectee *i* as well as the infector *j*. For a directed graph, *v*_*hij*_ equals 1 if an edge from the infector *j* to the infectee *i* in the household *h* is present or 0 if it is absent. The model for household transmission is

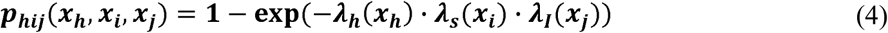

where *λ*_*h*_(*x*_*h*_), *λ*_*S*_(*x*_*i*_) and *λ*_*I*_(*x*_*j*_) characterize the general risk of household transmission in the household *h*, the susceptibility of the infectee *i* and the infectivity of the infector *j* respectively.

## 3 Methods

### 3.1 Random directed graph for TB transmission

There are important differences between the transmission dynamics of TB and influenza that impact the construction of the RDM. Unlike influenza, a large portion of individuals with latent TB infection remain in that state indefinitely and are not infectious. Those who progress to disease do so on a time scale of months or even years, compared to the rapid progression of influenza within days or hours of infection. It is generally accepted that LTBI is a noninfectious state, which may not be the case in influenza. The impact of this on the model is that in influenza, all infected cases are considered infectious, while in TB, only the LTBI cases who progress to active TB disease are infectious. To formalize our TB transmission model, we classify household contact study participants into three categories: 1): the active TB diseased (ATB) cases as the participants who are either the index cases or secondary TB diseased cases in household contact study; 2): the latent TB infection (LTBI) cases who have a TST reading ≥ 10 mm recorded by household contact study; and 3): the no TB infection (NTBI) cases whose TST reading(s) is constantly below 10 mm. The outcome of our model is infection status which is 1 for the ATB and LTBI cases and 0 for the NTBI cases.

An RDM for TB requires the following information: 1) observed infection status for all household members, namely ATB, LTBI, and NTBI; 2) dates of diagnoses for all ATB cases; and 3) covariate information on the individuals and environment that correlates with transmission.

The graph is drawn by adhering to three rules, modifying the rules in 2.2 to be specific to TB. These are:

1. Only ATB cases can send edges and they can only send edges to others within the same household.
2. ATB cases can only send edges to other ATB cases with later diagnosis dates and LTBI cases, as an ATB case can only infect those who progress to ATB at a later date and LTBI cases.
3. To produce a graph that is consistent with the observed data, the ATB case(s) with earliest diagnosing date must receive an edge from outside the household. However, we do not consider the edge of the earliest ATB cases for our estimation and results when community controls are not enrolled since including these edges in a household contact study without community controls would upwardly bias the estimates of extra-household transmission force.
4. ATB and LTBI case(s) should receive at least one edge and NTBI case(s) should receive no edges.

An example graph for TB is shown in Figure 1. There are three main assumptions in this paper. First, we assume the diagnosis dates and (unobserved) infection dates have the same temporal ordering, so that we can use them to determine infectors and infectees. Second, we assume households as well as edges are independent, meaning the edges drawn in one household do not influence the edges drawn in another household. Third, extra-household edges come from another (unknown) individual who lives in the same community.

**Figure 1:**
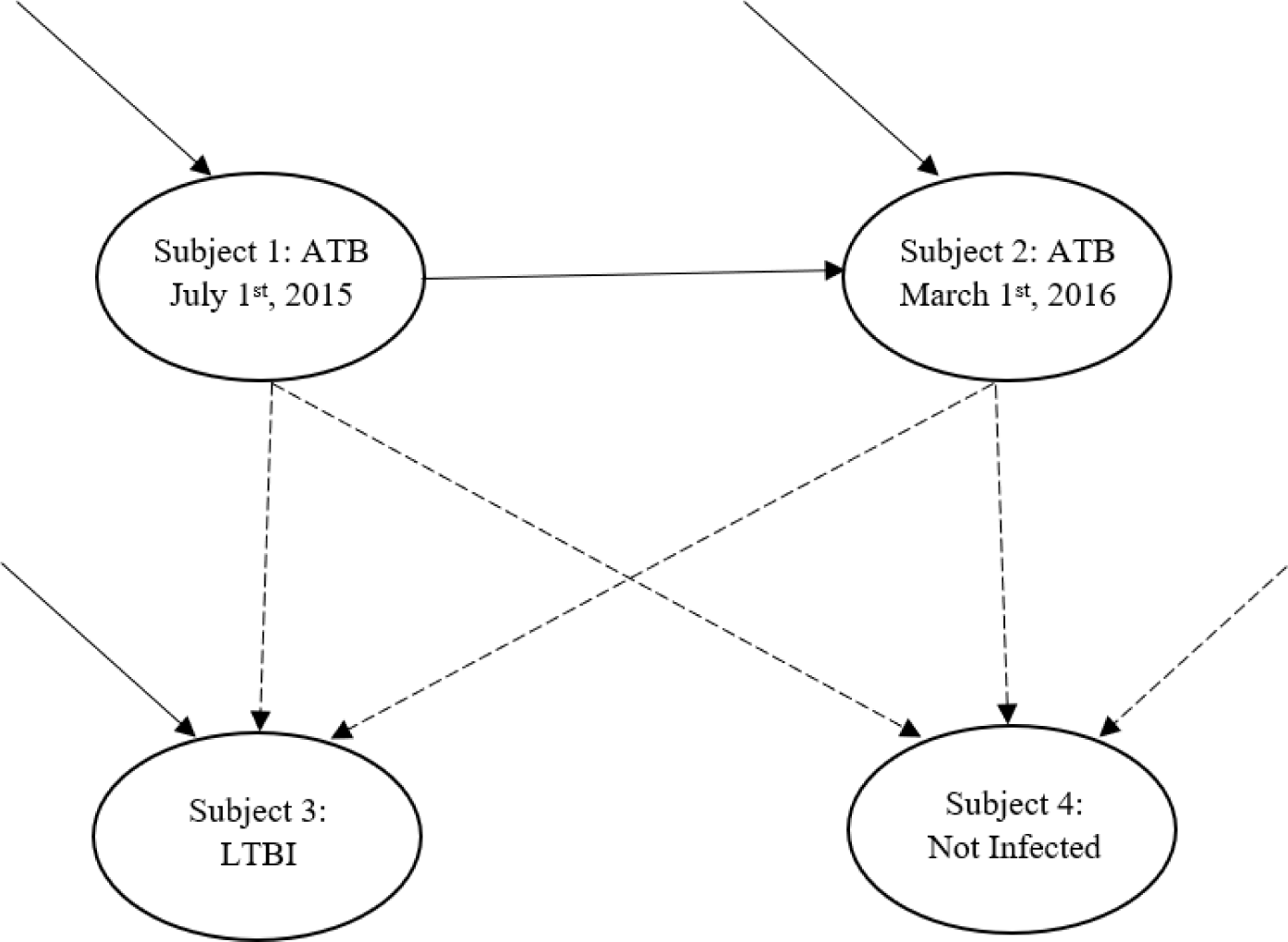
Example of a plausible directed graph for a household with 2 active TB cases (subject 1 and subject 2), 1 LTBI case (subject 3) and 1 non-infected (NTBI) case (subject 4). A solid arrow represents the presence of a potential transmission path and a dashed arrow represents the absence of a potential transmission path. Subject 1 is the ATB case with earliest diagnosing date (July 1^st^, 2015) and he/she should only receive an edge from outside the household. This edge is not considered when community controls are unavailable. Subject 2 is an ATB case but later than subject 1, so he/she could receive an edge from subject 1 and/or from outside the household (in this graph, he/she receives both edges). Subject 3 is an LTBI case and should at least receive one edge (in this graph, the edge is from outside the household), but he/she cannot send edges to someone else. Subject 4 is not infected, so he/she does not receive any edges.

### 3.2 TB transmission model with powerful predictors

In general, the likelihood function of a graph for TB is modified based on the likelihood function (2):

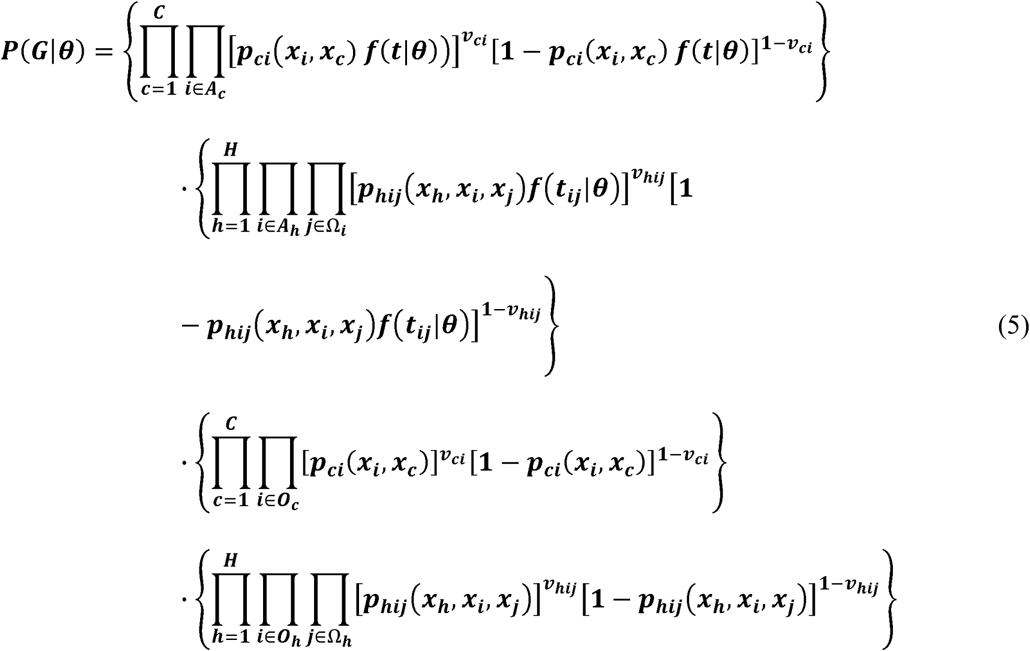

where *A*_*C*_ and *A*_*h*_ refer to the collection of all ATB cases (except the first in each household) in the community *c* and in the household *h* respectively. Similarly, *O*_*C*_ and *O*_*h*_ refers to the collection of all non-ATB cases in the community *c* and in the household *h* respectively. Ω_*i*_ is the collection of infectors that could infect the infectee *i* and it is a subset of Ω_*h*_. For example, Ω_*2*_ in Figure 1 is {subject 1, community} and Ω_*h*_ is {subject 1, subject 2, community}. We modified the likelihood function for ATB cases specifically: *f* is the density function of serial interval and *t*_*ij*_ is the lag between the dates of diagnoses of infector *j* and infectee *i*, i.e., *t*_*ij*_ *= d*_*j*_ *-d*_*i*_. t is a random number drawn from *f*(*t*|*0*) and represents the serial interval between the infectee *i* and an unknown infector outside the household. The reason for such modification is to incorporate information about the serial interval *f* and dates of diagnoses *d*_*i*_, *d*_*j*_ that are only available for ATB cases into the calculation of the relative likelihood of household transmission to extra-household transmission, with the hope that we can better impute the transmission paths between a pair of ATB cases.

In general, when one has enough powerful predictors for imputing transmission paths, the model for extra-household transmission and household transmission follows from (3) and (4):

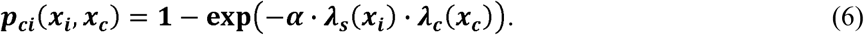

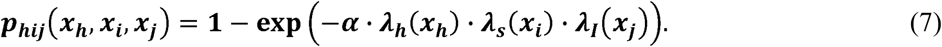

For the model above, α characterizes the risk of TB infection for the baseline group and all the predictors (i.e., *x*_*i*_, *x*_*C*_, *x*_*h*_, *x*_*j*_) are dummy variables representing risk factors for TB infection. Therefore, the baseline group in this case refers to the group of people who have the minimum risk of TB infection when the most important risk factors of TB infection have been controlled for in the transmission model. In this case, we think the risk of household transmission should be approximately the same as the risk of extra-household transmission for the baseline group. The model parameterization above can solve the non-identifiability issue by itself, and therefore it is estimable for a household contact study without community controls. However, this is only appropriate if most powerful predictors for TB transmission have been included in the model, an assumption that is impossible to test in practice and likely not achieved.

### 3.3 TB transmission model with insufficient predictors

If we do not have enough powerful predictors for describing TB transmission (due to measurement error, poor diagnostics, etc.), the earlier model is no longer appropriate as the baseline risks of TB infection for household and different communities will not be identical. Therefore, we need to reparameterize the transmission model (6) and (7)

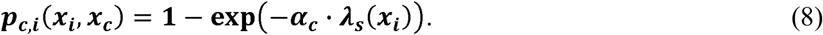

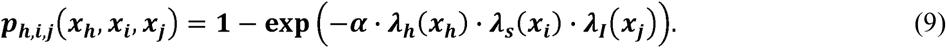

In this model, *α*_*c*_ characterizes the baseline risk of extra-household transmission in the community *c* (*c* = 1,…, *c*) and *a* characterizes the baseline risk of household transmission. The above model parameterization is non-identifiable and thus non-estimable for a household contact study without community controls, even after taking dates of diagnoses for ATB cases into account. This is because the majority of infectees are typically LTBI cases and information on the dates of diagnosis is not available, making it challenging to impute transmission paths based on insufficient predictors. Consequently, the household contact study does not provide any direct information on transmission outside of enrolled households, if both community controls and powerful predictors are unavailable.^17^

To address this issue, there are two strategies: The first strategy is to enroll community controls, which are used to identify extra-household transmission and thus separate household transmission from extra-household transmission. The drawback of this strategy is a large number of community controls may not be available in practice. The second strategy is to modify the MCMC algorithm for the household contact study data without community controls by a priori weighting extra-household transmission paths versus household transmission paths (or vice versa), which is detailed next.

### 3.4 Bayesian inference

We introduce a new MCMC algorithm that uses random weights to impute the transmission paths between an ATB case and an LTBI case. This approach allows researchers to incorporate knowledge about the relative strength of extra-household to household transmission when the model structure does not easily permit this information to be included as a proper prior. The weighted MCMC algorithm has the following steps:

1. Choose a lognormal distribution with the mean μ and the standard deviation σ from which to draw the weights.
2. In each iteration, draw a random number *t* from the density function of serial interval and a random number *r* from the lognormal distribution.
3. Update the parameters with the following probability of acceptance:

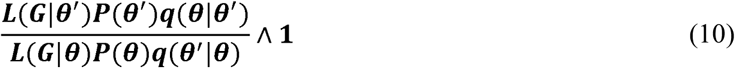

based on the likelihood function *L*(*G*|*θ*), the prior *P*(*θ*) and the proposal *q*(*θ* ′| (*θ*) for the current value *θ* and the candidate *θ* ′.
4. Update the graph for each infected individual, except the earliest ATB case in each household, with the probability of acceptance for ATB and LTBI defined separately.

### 4.1. For an ATB recipient *i* the acceptance probability is

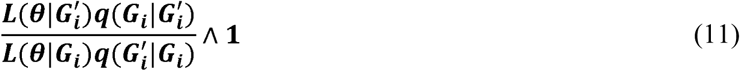

where *G*_*i*_ is a graph that shows presence and absence of all edges leading to the infectee *i*. The new graph 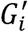 is obtained by randomly deleting or adding one path based on *G*_*i*_ The proposal ratio is:

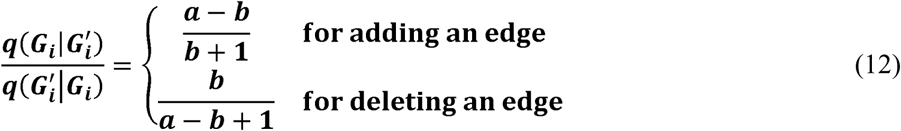

where *a* is the total number of possible edges leading to individual *i* and *b* is the number of edges in.*Gi*^15^

### 4.2. For a LTBI recipient *i* the acceptance probability is

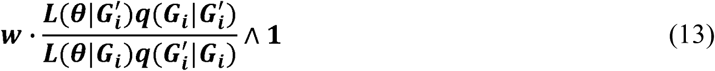

where *w* is determined as follows based on the weights. If μ < 0, household transmission paths would be penalized for each LTBI infectee by defining the weights by

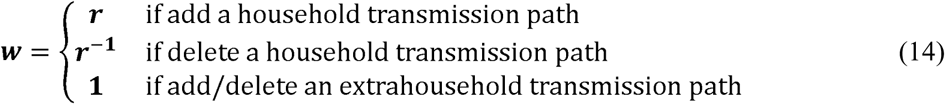

If μ > 0, extra-household transmission paths would be penalized for each LTBI infectee by defining the weights by

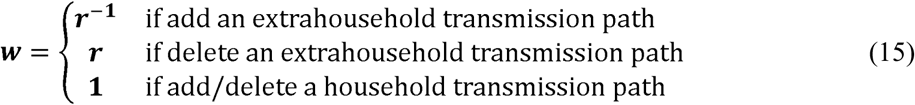

When either powerful predictors or community controls are available for the household contact study, weights are not needed and the above algorithm can be simplified (the simple MCMC algorithm). In each iteration,

1. Draw a random number t from the density function of serial interval
2. Update the parameters with the following probability of acceptance:

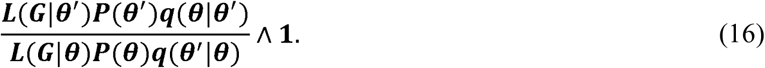
3. Update the graph for each infected individual, except the earliest ATB case in each household, with the following probability of acceptance:

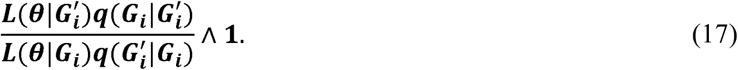

### 3.5 DIC and its application

When strong prior knowledge about *w* is not available, it is recommended that the MCMC algorithm is repeatedly run with multiple distinct weighting schemes, i.e., lognormal distributions with different μ. The idea is to repeatedly draw posterior samples under various weighting schemes ranging from weights strongly favoring household transmission to weights strongly favoring extra-household transmission and select the best-fitting weighting scheme(s) among them. In this section, we discuss how to use DIC to compare multiple samples drawn from the joint posterior with distinct weighting schemes.

*P*(*y*| *θ*) can be approximated in simulation by

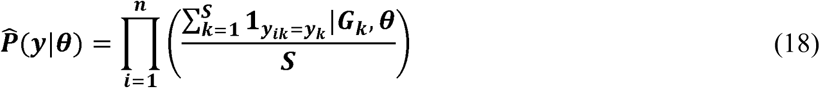

where S is the number of iterations for simulation and *G*_*k*_ is the observed graph in each iteration. The DIC is computed based on 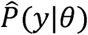, and in general we select weighting schemes to avoid under-weighting such that the difference between the DIC of selected weighting scheme and the minimum DIC should be no larger than 5. We also want to avoid over-weighting by selecting weighting scheme(s) that should not be more informative (i.e., μ should not be further away from 0) than the one with minimum DIC. Selected weighting scheme(s) should be consistent with one’s knowledge about whether extra-household transmission is stronger than household transmission (or vice versa). We illustrate the application of DIC in the results section.

## 4 Results

### 4.1 Simulation 1: Simulation with powerful predictors

We simulated TB epidemics for 1500 households with the following dichotomous predictors: age (</≥ 18), biological sex, household crowding (number of inhabitants per room > 2), sleeping proximity (sleeping in the same room with an ATB case), majority of time at home, severity of disease for ATB cases, high community burden, high socioeconomic status, and proximity to someone with a cough (Table 1). We assumed transmission was fully determined by the predictors above:

**Table 1:** Simulation results for model with powerful predictors.

| Baseline and relative susceptibility parameters | Simulation value | Mean of Median | Number of times 95% C.I covers the simulation value in 1000 simulations |
| --- | --- | --- | --- |
| $\alpha$: baseline risk | 0.03 | 0.03 | 965 |
| $\lambda_s^{adult=1}$: adult vs. child | 1.2 | 1.21 | 978 |
| $\lambda_s^{female=1}$: female vs. male | 1 | 1 | 988 |
| $\lambda_h^{crowd=1}$: crowded vs. uncrowded household | 1.5 | 1.5 | 952 |
| $\lambda_s^{sleep=1}$: slept with vs. not slept with TB patient | 5.5 | 5.02 | 943 |
| $\lambda_s^{time=1}$: spent more time vs. less time in household | 2 | 1.92 | 956 |
| $\lambda_I^{severity=1}$: Infector with high severity vs. low severity | 2.8 | 2.62 | 966 |
| $\lambda_c^{burden=1}$: lived in high vs. low burden community | 3.5 | 3.26 | 970 |
| $\lambda_c^{ses=1}$: lived in higher vs. lower socio-economical community | 3 | 2.86 | 977 |
| $\lambda_s^{cough=1}$: experiencing cough vs. no such experience in community | 2.5 | 2.44 | 971 |

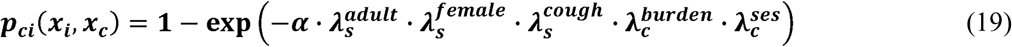

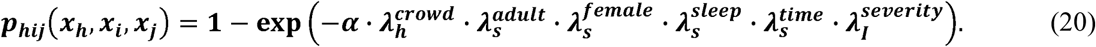

where 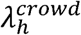 equals 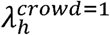 for a crowded household (crowd = 1) or 1 for a non-crowded household (crowd = 0). The interpretations for all other parameters are similar.

We only selected the households with at least one ATB case for subsequent estimation and, in essence create a “perfect” household contact study where we assumed we were able to trace and enroll all TB patients and their household contacts. The size of the final data was similar to the Brazilian household contact study (average number of TB households was 155). The simulation generated 1000 simulated datasets and for each dataset the simple MCMC algorithm was iterated 220,000 times with a burn-in of 20,000 and a thinning of 20. Each run took about 2 hours by using the Rcpp package on a laptop (Intel® core i7-8550U CPU@1.8 GHz).

The results showed that the simple MCMC algorithm generated good estimates and reliable confidence intervals for all parameters, without using any weights or community controls (Figure 2). The actual coverage rates ranged from 94.3% to 98.8% across the parameters (Table 1). Therefore, for household contact study data (i.e., the households with at least one ATB case; henceforth we refer it as the HHC data) with powerful predictors, we confirm that there was no need to employ community controls or weights and there is no concern regarding the non-identifiability issue.

**Figure 2:**
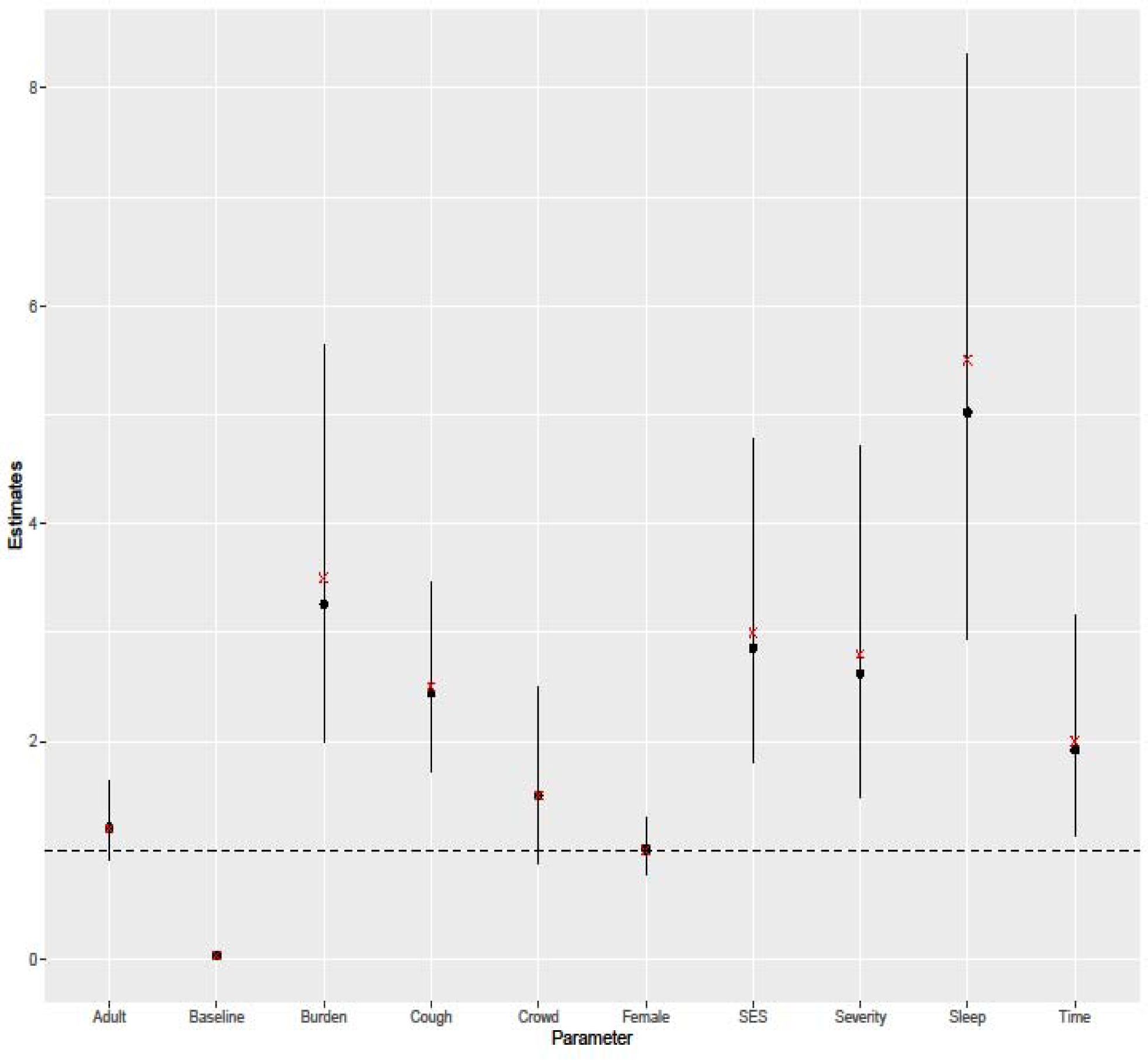
Simulation results for the model with powerful predictors. The results were summarized over 1000 simulated datasets which were similar to Brazilian household contact study. The solid lines represent the average 95% C.I. for all model parameters, and the black dots represent the mean of medians in the posterior sample of parameters. The red crosses are the simulated values of the parameters. The horizontal dashed line corresponds to the value 1, which is the threshold of whether a predictor signals higher risk of TB infection. It is clear that for the model with powerful predictors the MCMC algortihm (without weights) can lead to good estimates and good identifiability, even without any community controls.

### 4.2 Simulation 2: Simulation with insufficient predictors

As previously discussed, in practice we likely do not have a set of predictors that are sufficient to estimate the transmission paths of the RDM. In this simulation, we only used three predictors (which have been defined in the first simulation): age (</≥ 18), biological sex, and household crowding. We assumed transmission was fully determined by the predictors above:

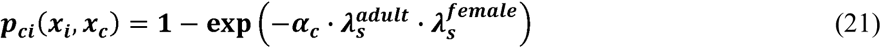

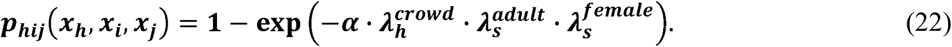

We consider four scenarios depicting varying relative magnitudes of household transmission.

1. Extra-household transmission accounted for approximately 90% of the total infections.
2. Extra-household transmission accounted for 50-60% of the total infections.
3. Household transmission accounted for 50-60% of the total infections.
4. Household transmission accounted for 90% of the total infections.

We simulated 25 datasets for each scenario and for each dataset we ran three different kinds of analyses. First, we used all households, including those with and without ATB case(s), and ran the simple MCMC algorithm on this data (the whole data approach). Second we used the HHC data (only households with at least one ATB case) and ran the simple MCMC algorithm on it. Third, we used the HHC data and separately ran the weighted MCMC algorithm with 10 different weighting schemes. All MCMC algorithms were iterated 220,000 times with a burn-in of 20,000 and a thinning of 20. The final size of the HHC data was similar to the Brazilian household contact study (the average number of TB households was 177, with minimum = 140 and maximum = 214).

We chose five of the ten weighting schemes to favor extra-household transmission and the other five to favor household transmission and show how DIC can be used to select the best weights. For the weighting schemes favoring extra-household transmission, we chose the mean of lognormal distribution as −0.25, −0.2, −0.15, −0.1, and −0.05 with standard deviation as 0.05. For the weighting schemes favoring household transmission, the mean of lognormal distribution was −0.02, −0.04, −0.06, −0.08 and −0.1 with standard deviation 0.02. The average DIC of the 10 posterior samples drawn under the weighting schemes in the four simulation scenarios are shown in Figure 3. DIC correctly informs us about whether a weighting scheme should favor extra-household transmission (or household transmission) under the simulation scenario 1, 2 and 4. For example, in scenario 1 where weighting scheme should strongly favor extra-household transmission, the weighting schemes favoring household transmission all had significant higher DIC than the weighting schemes favoring extra-household transmission, suggesting that the weighting scheme(s) favoring household transmission poorly fit to the data. In scenario 3, we still observed that the weighting schemes favoring household transmission have lower DIC than the weighting schemes favoring extra-household transmission, although the differences were not statistically significant. Not surprisingly, DIC is more informative under the scenarios 1 and 4 than under the scenarios 2 and 3. However in scenarios 2 and 3 it is not clear how consequential the weights are since household and extra-household transmission are almost equally likely.

**Figure 3:**
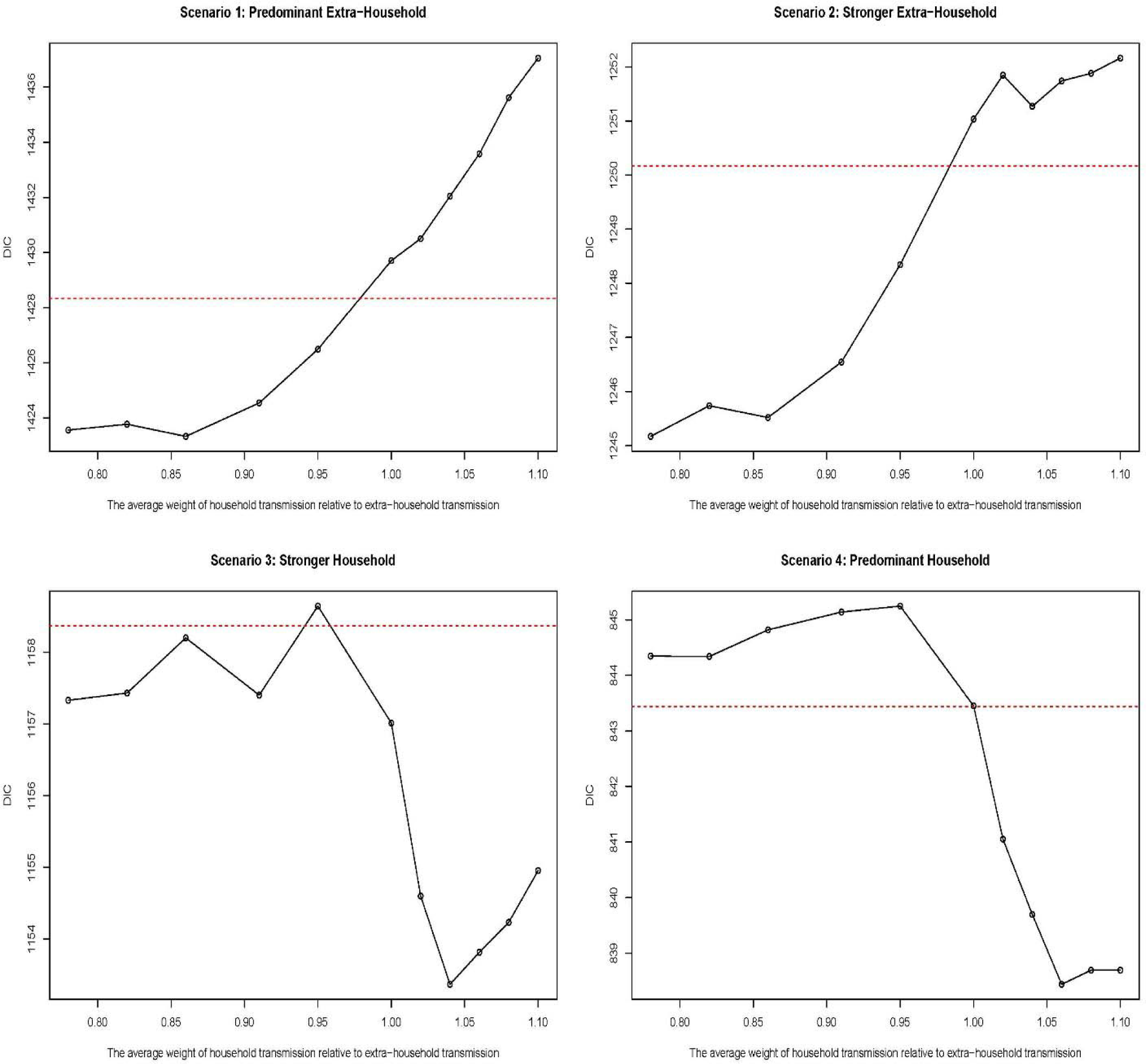
The plot of average DIC for 11 posterior samples, 10 of which were drawn under the weighting schemes and one based on the simple MCMC algorithm (shown at weight = 1). The red dashed line represents the value which equals the minimum DIC + 5, which is the threshold for deciding whether a weighting scheme is significantly different from the minimum DIC. In this case, we only consider the weighting schemes that have DIC lower than this threshold.

Next, we compared the parameter estimates based on the HHC data without weights, the whole data without weights, and the HHC data with weights. For the third approach, we selected weighting schemes using DIC. Using weights on the HHC data helped identify the extra-household and household baseline transmission forces and thus reduce bias of the estimates, compared to the estimates based on the HHC data without weights (Figure 4). Using community controls helped identify the baseline extra-household transmission forces and improved the accuracy of estimates compared to the other two approaches. When household transmission is stronger, community controls also automatically leaded to identification of the baseline household transmission force (Tables 4 and 5). However, when extra-household transmission dominates, community controls do not appear sufficient to accurately estimate household baseline transmission (Tables 2 and 3).

**Figure 4:**
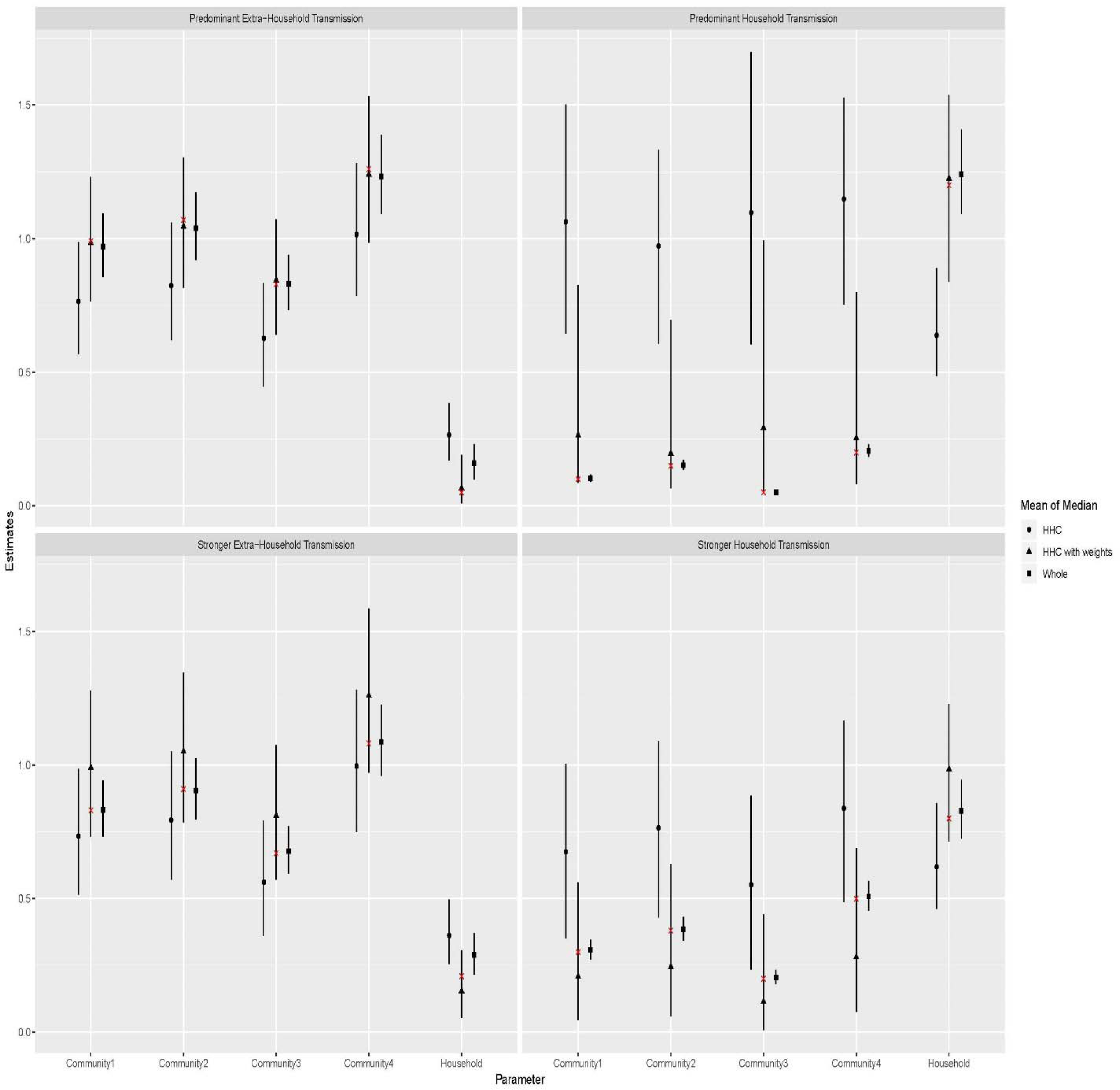
Comparing the baseline transmission force estimates of household and the communities across the four simulation scenarios. The line represents the average 95% CI for 25 datasets; The black symbols represent mean of median estimates among 25 datasets, and the red cross represent the underlying simulation values. ‘HHC’ in the figure above refers to the approach that we used the HHC data without weights.

**Table 2:** Simulation result for scenario 1: extra-household transmission is predominant. The results are described in the following format: mean of median (the number of times that CI covers the simulation value in 25 datasets). For example, 0.16 (3) means the average median estimate was 0.16 for 25 datasets and confidence interval covered the simulation value 3 times for 25 datasets.

| Baseline and relative susceptibility parameters | Simulation value | Estimates based on the whole data without weights | Estimates based on the household contact study without weights | Estimates based on the household contact study with weights |
| --- | --- | --- | --- | --- |
| $\alpha$: baseline household | 0.05 | 0.16 (3) | 0.27 (0) | 0.07 (24) |
| $\alpha_{c1}$: baseline for the first community | 0.99 | 0.97 (24) | 0.76 (13) | 0.98 (25) |
| $\alpha_{c2}$: baseline for the second community | 1.07 | 1.04 (25) | 0.82 (11) | 1.05 (25) |
| $\alpha_{c3}$: baseline for the third community | 0.83 | 0.83 (22) | 0.63 (14) | 0.84 (21) |
| $\alpha_{c4}$: baseline for the fourth community | 1.26 | 1.23 (24) | 1.02 (13) | 1.24 (23) |
| $\lambda_s^{crowd=1}$: crowded vs. non-crowded household | 1.2 | 1.07 (23) | 1.04 (22) | 0.97 (24) |
| $\lambda_s^{adult=1}$: adult vs. child | 1.2 | 1.2 (25) | 1.2 (25) | 1.2 (25) |
| $\lambda_s^{female=1}$: female vs. male | 1 | 0.99 (21) | 0.98 (22) | 0.98 (23) |

**Table 3:**
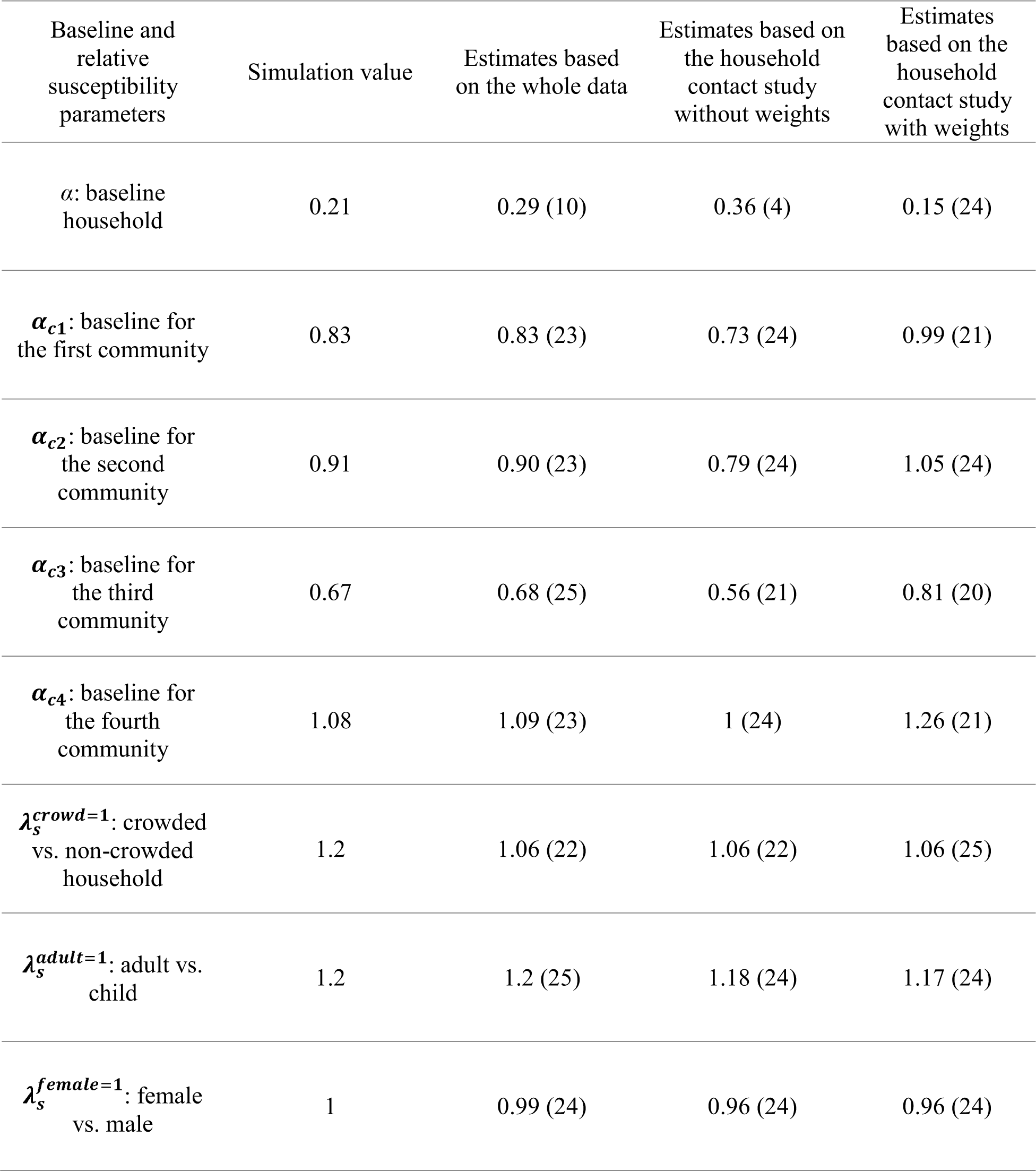
Simulation result for scenario 2: extra-household transmission is stronger. The results are described in the following format: mean of median (the number of times that CI covers the simulation value in 25 datasets).

**Table 4:** Simulation result for scenario 3: household transmission is stronger. The results are described in the following format: mean of median (the number of times that CI covers the simulation value in 25 datasets).

| Baseline and relative susceptibility parameters | Simulation value | Estimates based on the whole data | Estimates based on the household contact study without weights | Estimates based on the household contact study with weights |
| --- | --- | --- | --- | --- |
| $\alpha$: baseline household | 0.8 | 0.83 (23) | 0.62 (12) | 0.98 (15) |
| $\alpha_{c1}$: baseline for the first community | 0.3 | 0.31 (22) | 0.67 (10) | 0.21 (19) |
| $\alpha_{c2}$: baseline for the second community | 0.38 | 0.38 (25) | 0.76 (7) | 0.24 (20) |
| $\alpha_{c3}$: baseline for the third community | 0.2 | 0.21 (20) | 0.55 (13) | 0.11 (24) |
| $\alpha_{c4}$: baseline for the fourth community | 0.5 | 0.51 (23) | 0.84 (10) | 0.28 (17) |
| $\lambda_s^{crowd=1}$: crowded vs. non-crowded household | 1.2 | 1.14 (23) | 1.16 (24) | 1.13 (24) |
| $\lambda_s^{adult=1}$: adult vs. child | 1.2 | 1.18 (23) | 1.15 (23) | 1.16 (23) |
| $\lambda_s^{female=1}$: female vs. male | 1 | 1 (24) | 1 (23) | 1 (23) |

**Table 5:** Simulation result for scenario 4: household transmission is predominant. The results are described in the following format: mean of median (the number of times that CI covers the simulation value in 25 datasets).

| Baseline and relative susceptibility parameters | Simulation value | Estimates based on the whole data | Estimates based on the household contact study without weights | Estimates based on the household contact study with weights |
| --- | --- | --- | --- | --- |
| $\alpha$: baseline household | 1.2 | 1.24 (22) | 0.64 (4) | 1.23 (20) |
| $\alpha_{c1}$: baseline for the first community | 0.1 | 0.1 (24) | 1.06 (1) | 0.26 (22) |
| $\alpha_{c2}$: baseline for the second community | 0.15 | 0.15 (24) | 0.97 (4) | 0.2 (21) |
| $\alpha_{c3}$: baseline for the third community | 0.05 | 0.05 (24) | 1.1 (2) | 0.29 (22) |
| $\alpha_{c4}$: baseline for the fourth community | 0.2 | 0.21 (20) | 1.15 (4) | 0.25 (20) |
| $\lambda_s^{crowd=1}$: crowded vs. non-crowded household | 1.2 | 1.15 (24) | 1.15 (25) | 1.15 (24) |
| $\lambda_s^{adult=1}$: adult vs. child | 1.2 | 1.17 (22) | 1.1 (21) | 1.11 (23) |
| $\lambda_s^{female=1}$: female vs. male | 1 | 1.01 (24) | 1 (25) | 1 (25) |

Interestingly, the estimates of risk of infection for the covariates (age, crowding and biological sex) were comparable for all three estimation approaches (Figure 5). This indicates that the non-identifiability issue only exists for baseline extra-household and household transmission force estimates and does not appear to impact estimation of risk factors of TB transmission model when using RDMs.

**Figure 5:**
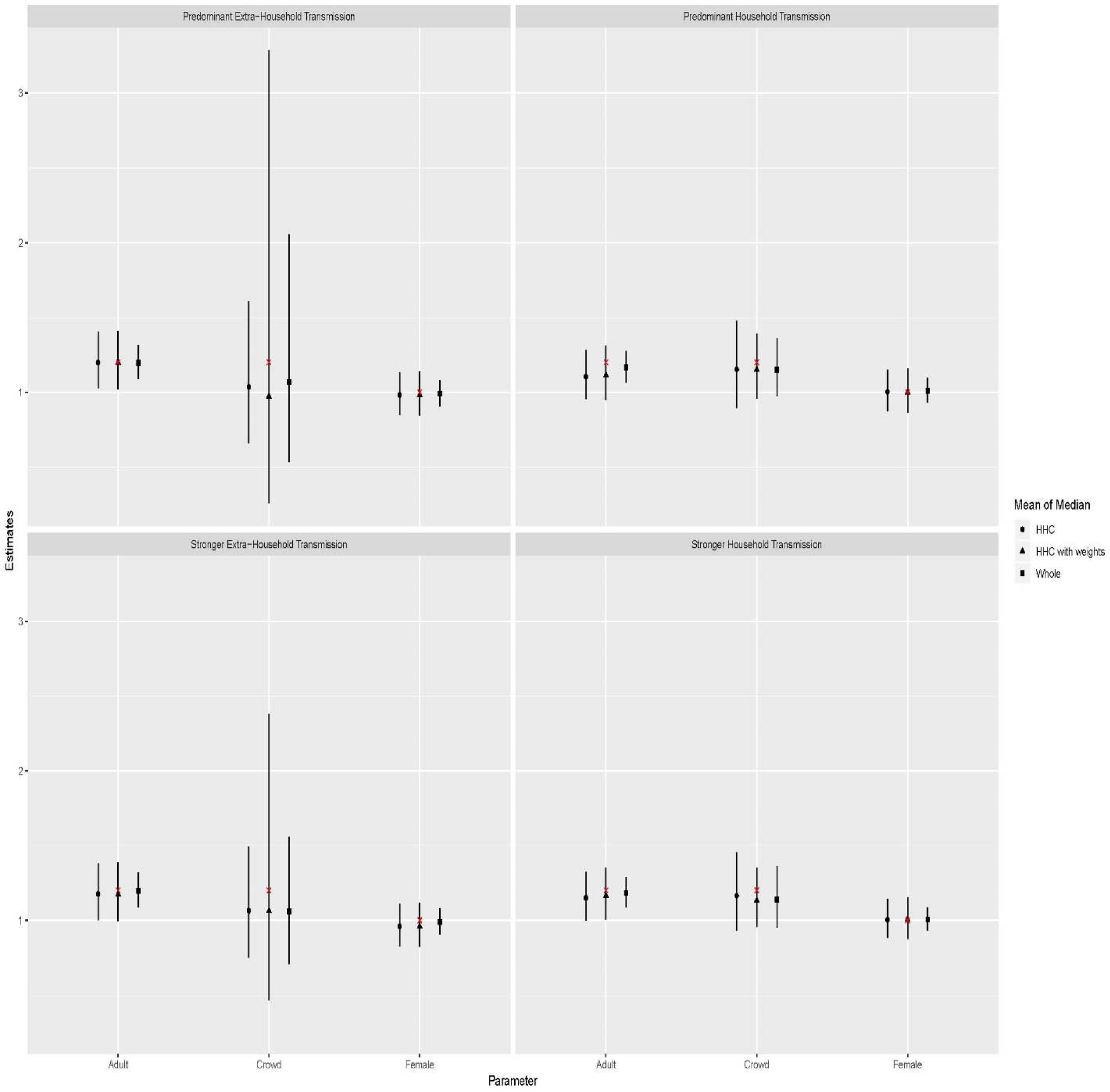
Comparing the risk factor estimates across the four simulation scenarios. The line represents the average 95% CI for 25 datasets; The black symbols represent mean of median estimates among 25 datasets, and the red cross represent the underlying simulation values. ‘HHC’ in the figure above refers to the approach that we used the HHC data without weights.

### 4.3 The Brazilian household contact study

The Brazilian household contact study enrolled households from four municipalities and the following predictors were used in the transmission model: adult (</ ≥ 18 years), biological sex, household crowding (number of inhabitants per room > 2). The first two variables were included in both the extra-household and household transmission model and the last one was used in the household transmission model only. Since Brazil has substantial TB in the community based on prior work, it is reasonable to assume that extra-household transmission is stronger than household transmission.^4,12^ To be objective in implementing the weighted MCMC algorithm we explore weights similar to those we used in the simulations (i.e. five favoring extra household and five favoring household transmission with standard deviation fixed at 0.05). All MCMC algorithms were iterated 220,000 times with a burn-in of 20,000 and a thinning of 20. The DIC suggests extra-household transmission is stronger than household transmission. (Figure 6) We then selected three weighting schemes whose μ (mean of lognormal distribution) were −0.2, −0.15, −0.1 since their DIC were not significantly different from the minimum DIC. We excluded the weighting scheme whose μ was −0.25 to avoid overweighting. Subsequently, we used the three corresponding posterior samples as parameter estimates. To enhance interpretability, we simulated TB epidemics based on the parameter estimates and reported the underlying TB transmission pattern as our result.^14,15^ In total, there were 30,000 simulated TB epidemics and each of them was done for 10,000 households in four municipalities, which were set to have similar features as the Brazilian household contact study.

**Figure 6:**
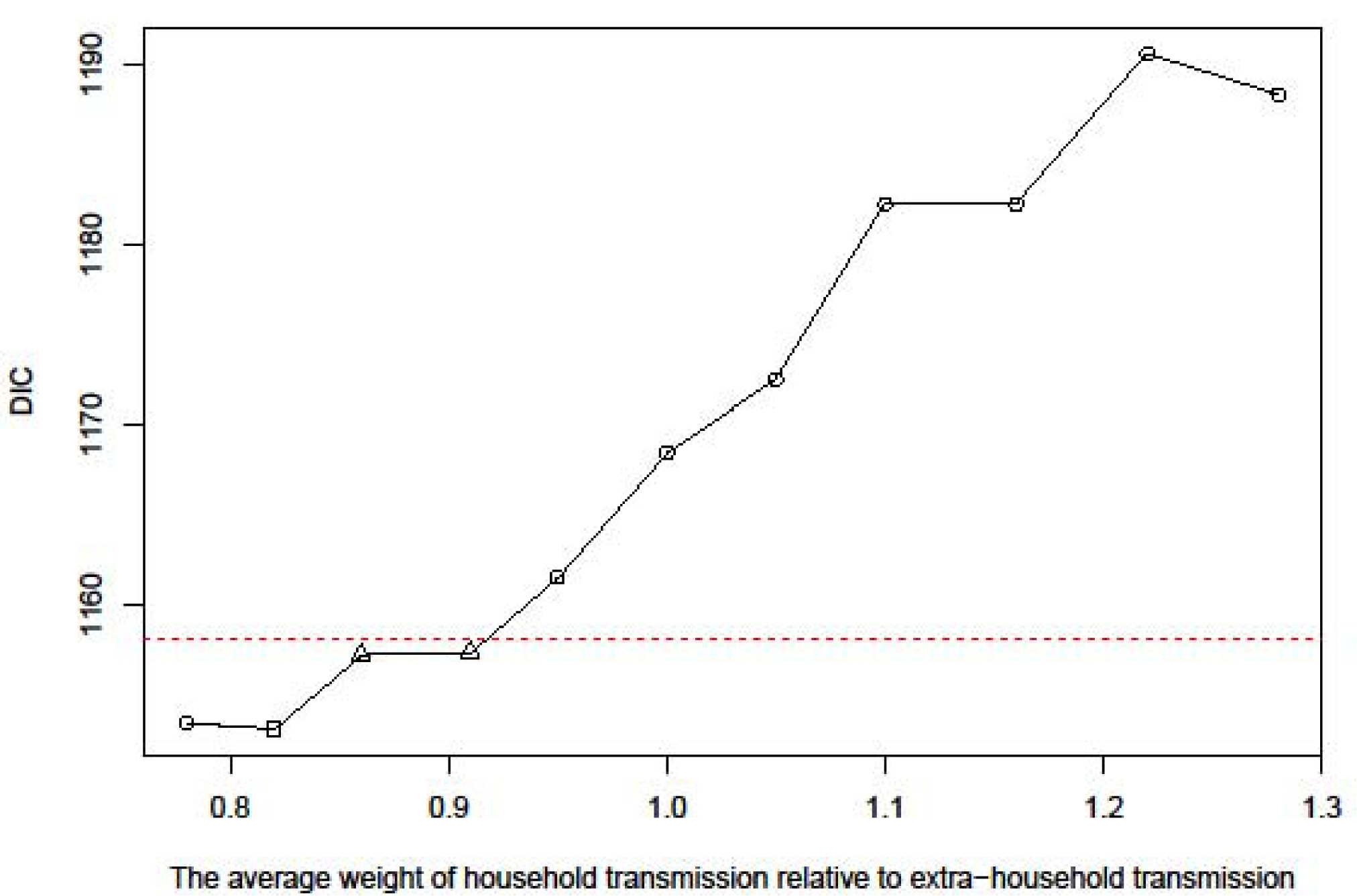
The plot of DIC for the Brazilian household contact study. We ploted DIC for 11 posterior samples, 10 of which were drawn under the weighting schemes and one based on the simple unweighted MCMC algorithm (x value = 1). The red dashed line represents the value which equals minimum DIC + 5, which is the threshold for deciding whether a weighting scheme is significantly different from the one with minimum DIC (represented by the square dot). In this case, we chose the three weighting schemes that represented by the square and triangle dots as our candidates.

Overall, the relative risk of being infected within the household to being infected outside the household was 0.23 (95% CI, 0.03-0.49), demonstrating that extra-household transmission was the predominant transmission force (Figure 7). Consistent with this, we estimate that 82% (95% CI, 63%-98%) of infections are attributed to extra-household transmisson while only 6% (95% CI, 1%-15%) of infections are estimated to be attributed to household transmission. Those proportions were consistent across the four municipalities (Figure 8). We also studied the risk factors for TB transmission (Figure 9) and found that adults had 9% more risk of TB infection than non-adults on average (RR: 1.09, 95% CI: 0.99-1.20). Females had the same of risk of TB infection compared to males (RR: 1.00, 95% CI: 0.91-1.10). Living in a crowded household did not increase the risk of TB infection (RR: 1.01, 95% CI: 0.99-1.04), likely due to the fact that crowding was only associated with household transmission which was minimal in this context.

**Figure 7:**
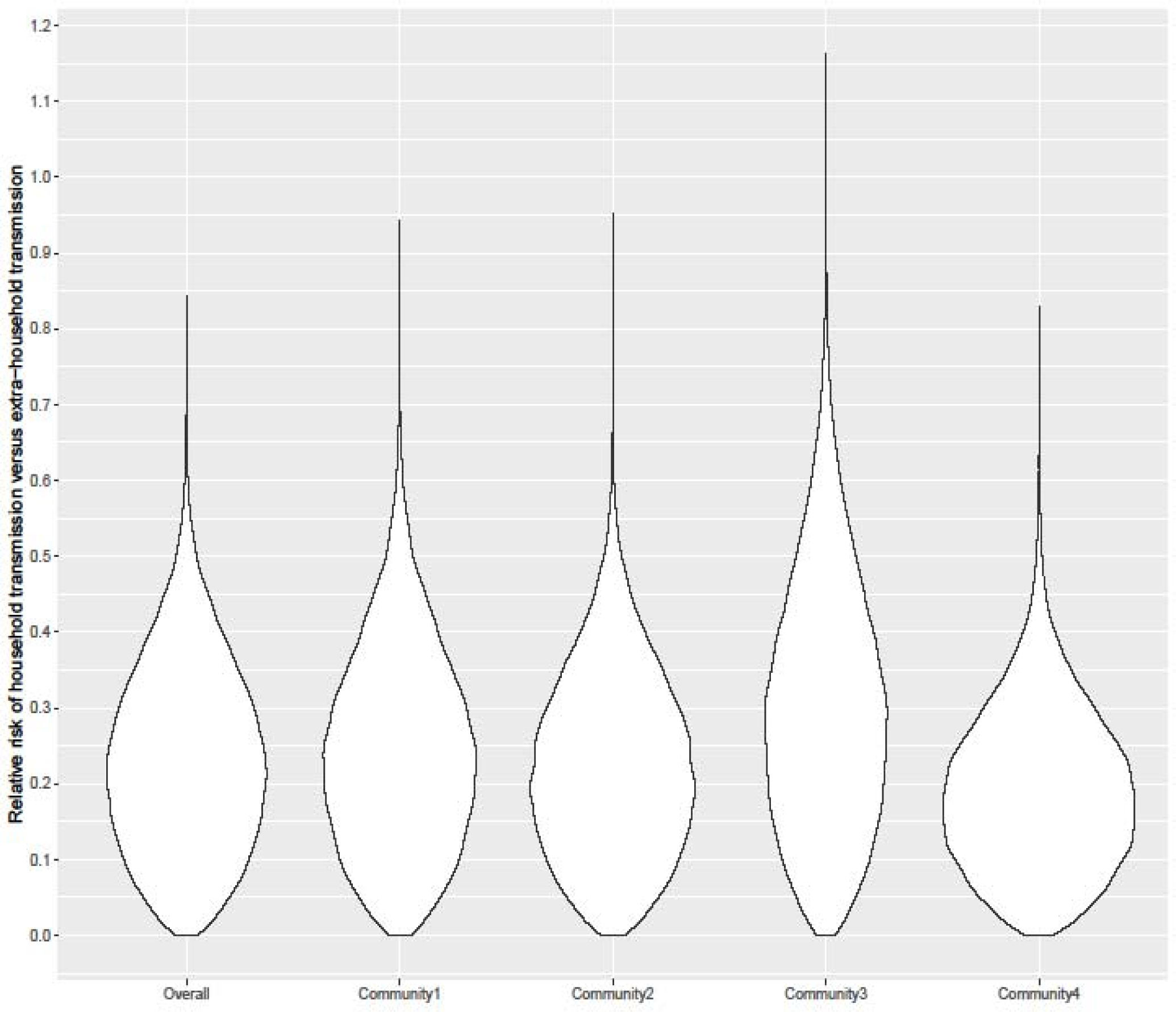
The violin plot of relative risk of household versus extra-household transmission across the four municipalities.

**Figure 8:**
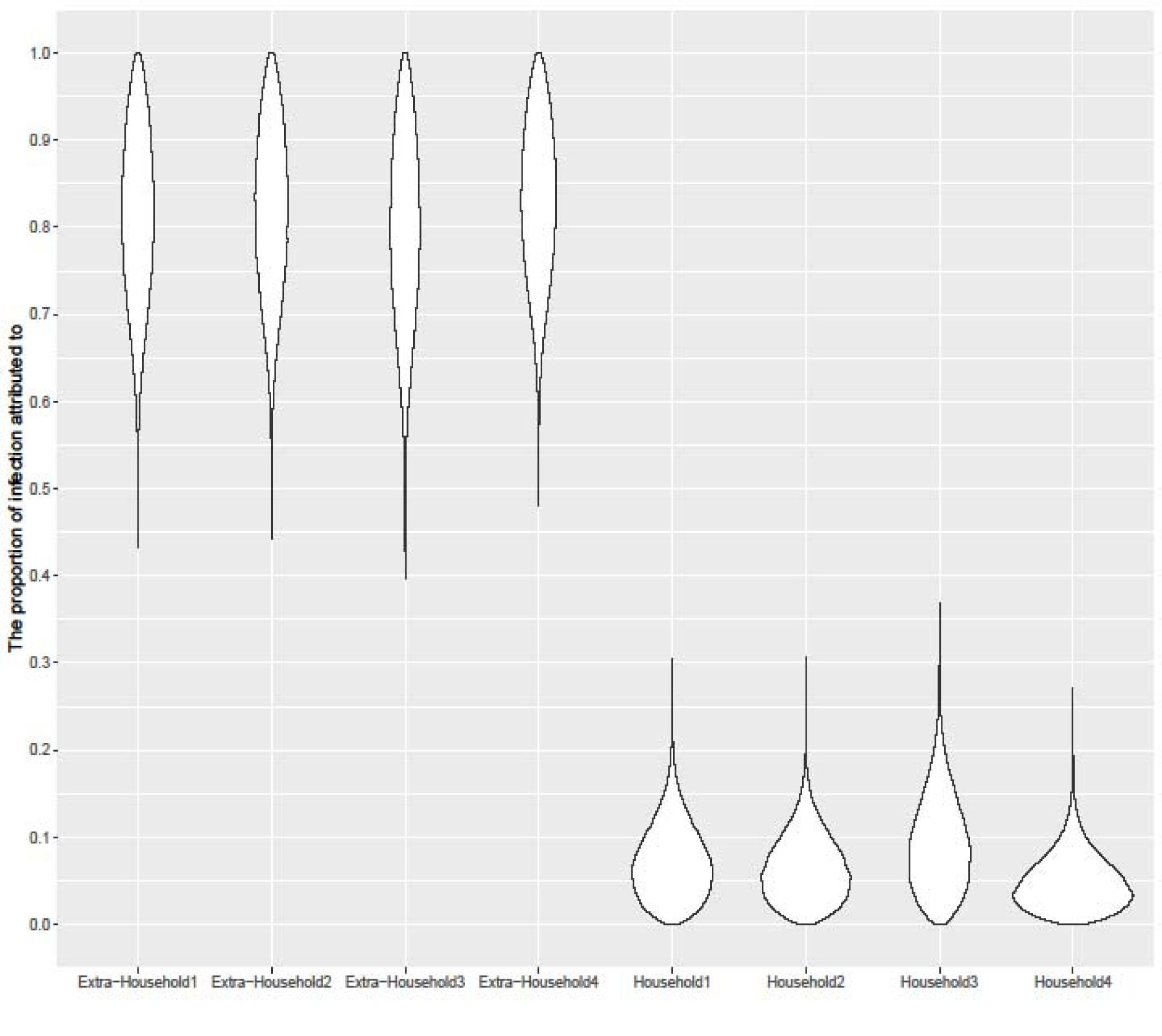
The violin plot of the proportions of TB infection attributed to extra-household and household transmission across the four municipalities.

**Figure 9:**
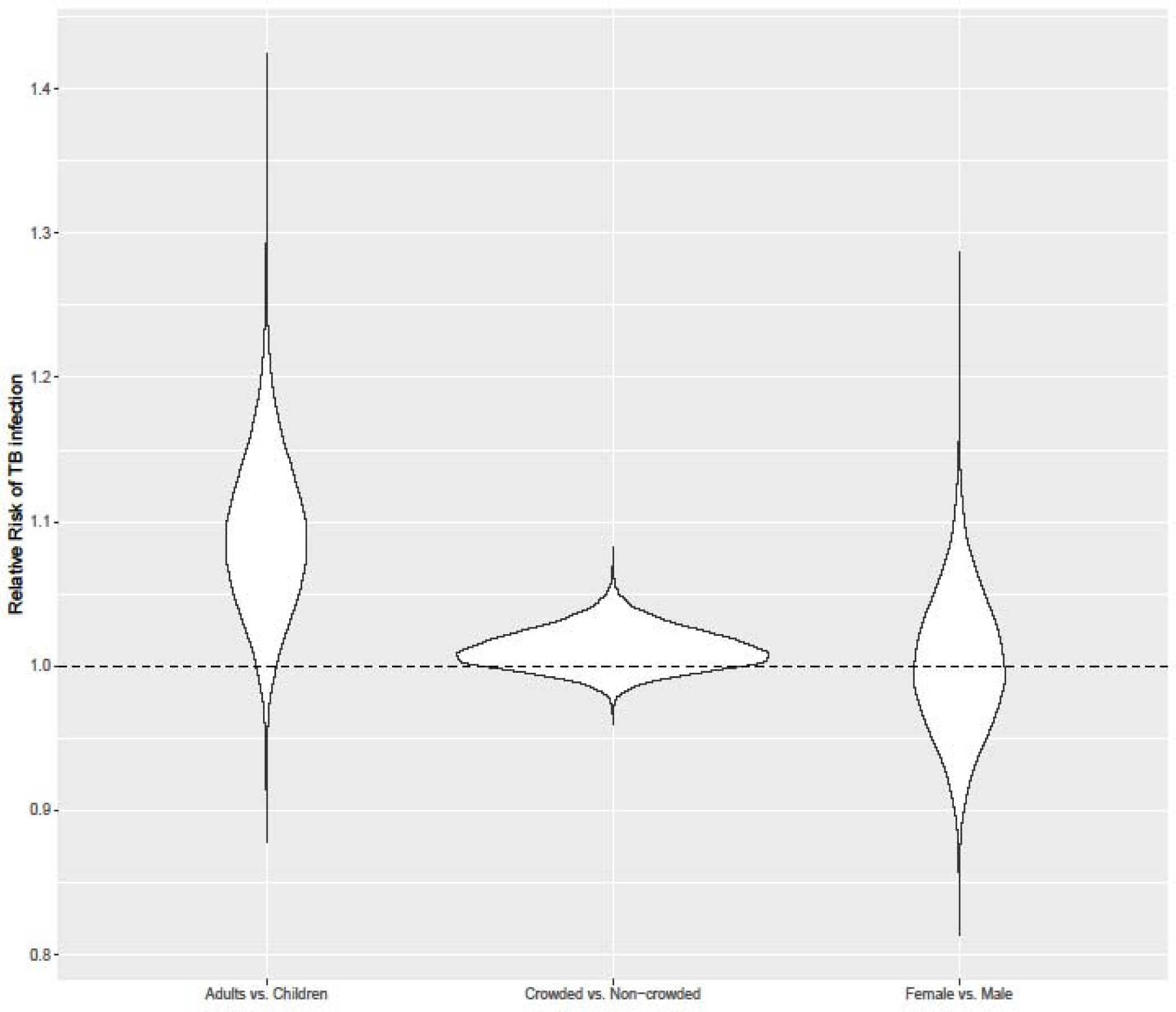
The violin plot of the relative risk of TB infection for the risk factors included in TB transmission model. The horizontal dashed line corresponds to RR = 1, which means a risk factor does not have any effect in TB transmission.

## 5 Discussion

In this study, we discuss the application of random directed graph model (RDM) to understand TB transmission. We first formalized rules of drawing random directed graph appropriate for TB transmission, and modified the likelihood model by treating ATB cases differently from LTBI and NTBI cases. By doing this, we can incorporate the diagnosing dates of ATB cases to better impute transmission paths among ATB cases as well as the temporal order of TB transmission chains. We further addressed the identification issue of RDM in this context with three different strategies: 1) control for the most powerful predictors in the TB transmission model; 2) enroll community controls in the household contact study; 3) use weights and DIC if the above two strategies become unviable. Having the identification issue addressed, simulation showed that the RDM could successfully estimate the relative risk and importance of household transmission versus extra-household transmission even without community controls. Furthermore, RDM is also a valuable tool for incorporating community controls and would gain power for both identification and estimation if they are available.^5^ RDM can consistently generate unbiased estimate of the relative risks of risk factors included in the transmission model, regardless of the usage of powerful predictors, community controls or weights. This is attractive compared to mixed model which is known to output biased estimates in the same setting.^4^

Our work is motivated by TB household contact study through which one seeks to infer household and extra-household transmission and their contributions to the general TB transmission dynamics. Traditionally, TB household contact studies assume TB transmission only happens within the household, which is not consistent with TB literature that suggests TB transmission in some settings is more likely to happen outside the household than in the household.^10,11^ Moreover, strain analysis using restriction fragment length polymorphism or spoligotyping of secondary diseased cases suggests that extra-household transmission could account for up to 70% of the total infections.^18-22^ However most of the models for household contact studies are logistic regression or mixed models assuming transmission only happens within the household.^8^ When using such models in a context where extra-household transmission is considerable, risk estimates would be biased especially for risk factors that solely associated with household transmission such as crowding, which may result in misguided public health planning.^4,23^ Therefore, it is critical to use models allowing concurrent estimation of household and extra-household transmission (such as UPM and RDM) in order to quantify the relative importance of household versus extra-household transmission.

There are limitations for our approach. First, our approach is simulation-based, which demands the model to be consistent with the simulation which should be accurate and realistic. In our paper, we assume the simulation is fully determined by controlled covariates in the model. However, this assumption is hardly testable and thus we don’t consider the confounding effect in this paper. Second, the transmission model parameters are not quite interpretable and to gain interpretability we recommend that the relative risk as well as risk estimates are obtained through simulation. Third, our approach is computationally intensive compared to other Bayesian approaches given the additional work on simulation and possibly weighting.

In summary, RDM is a valuable tool for TB household contact study as it provides additional knowledge on extra-household transmission as well as the roles of extra-household and household transmission in TB transmission dynamics. It worth emphasizing that, the knowledge gained by RDM cannot be picked up by a traditional statistical method, but it is clearly needed for public health programs to better plan their resources and target interventions.

## Data Availability

All data used in this manuscript is available online.

https://github.com/tenglongli/tb-rdm-method

## Supplementary Material

Supplemental material is available online and contains more details on the MCMC algorithm and simulation. The code and data for this paper are available at https://github.com/tenglongli/tb-rdm-method.

## Acknowledgements

We thank David Alland, Padmini Salgame, Jerrold Ellner and Reynaldo Dietze for sharing the data.

## References

1. WHO. Global tuberculosis report 2019. https://www.who.int/tb/publications/global_report/en/. Accessed: 2020-04-07.

2. WHO. The END TB Strategy. https://www.who.int/tb/strategy/end-tb/en/. Accessed: 2020-04-07.

3. Horsburgh CR Jr, Rubin EJ. Latent tuberculosis infection in the United States. New England Journal of Medicine. 2011; 364(15):1441–1448.

4. McIntosh AI, Doros G, Jones□López EC, Gaeddert M, Jenkins HE, Marques□Rodrigues P, Ellner JJ, Dietze R, White LF. Extensions to Bayesian generalized linear mixed effects models for household tuberculosis transmission. Statistics in Medicine. 2017; 36(16):2522–2532.

5. Whalen CC, Zalwango S, Chiunda A, Malone L, Eisenach K, Joloba M, Boom WH, Mugerwa R. Secondary attack rate of tuberculosis in urban households in Kampala, Uganda. PloS One. 2011;6(2).

6. Jones-López EC, Kim S, Fregona G, Marques-Rodrigues P, Hadad DJ, Molina LP, Vinhas S, Reilly N, Moine S, Chakravorty S, Gaeddert M. Importance of cough and M. tuberculosis strain type as risks for increased transmission within households. PloS One. 2014:9(7).

7. Hochberg NS, Sarkar S, Horsburgh Jr CR, Knudsen S, Pleskunas J, Sahu S, Kubiak RW, Govindarajan S, Salgame P, Lakshminarayanan S, Sivaprakasam A. Comorbidities in pulmonary tuberculosis cases in Puducherry and Tamil Nadu, India: Opportunities for intervention. PloS One. 2017;12(8).

8. Guwatudde D, Nakakeeto M, Jones-Lopez EC, Maganda A, Chiunda A, Mugerwa RD, Ellner JJ, Bukenya G, Whalen CC. Tuberculosis in household contacts of infectious cases in Kampala, Uganda. American Journal of Epidemiology. 2003;158(9):887–898.

9. Kilicaslan Z, Kiyan E, Kucuk C, Kumbetli S, Sarimurat N, Ozturk F, Yapici D, Al S, Erboran T, Iliksu N. Risk of active tuberculosis in adult household contacts of smear-positive pulmonary tuberculosis cases. The International Journal of Tuberculosis and Lung Disease. 2009;13(1):93–98.

10. Martinez L, Shen Y, Mupere E, Kizza A, Hill PC, Whalen CC. Transmission of Mycobacterium tuberculosis in households and the community: a systematic review and meta-analysis. American Journal of Epidemiology. 2017;185(12):1327–1339.

11. Martinez L, Lo NC, Cords O, Hill PC, Khan P, Hatherill M, Mandalakas A, Kay A, Croda J, Horsburgh CR, Zar HJ. Paediatric tuberculosis transmission outside the household: challenging historical paradigms to inform future public health strategies. The Lancet Respiratory Medicine. 2019;7:544–552.

12. McIntosh AI, Jenkins HE, Horsburgh CR, Jones-López EC, Whalen CC, Gaeddert M, Marques-Rodrigues P, Ellner JJ, Dietze R, White LF. Partitioning the risk of tuberculosis transmission in household contact studies. PloS One. 2019;14(10).

13. Demiris N, O’Neill PD. Bayesian inference for stochastic multitype epidemics in structured populations via random graphs. Journal of the Royal Statistical Society: Series B (Statistical Methodology). 2005;67(5):731–745.

14. Cauchemez S, Ferguson NM, Fox A, Mai LQ, Thanh LT, Thai PQ, Thoang DD, Duong TN, Hoa LN, Hien NT, Horby P. Determinants of influenza transmission in South East Asia: insights from a household cohort study in Vietnam. PloS Pathogens. 2014;10(8).

15. Tsang TK, Fang VJ, Ip DK, Perera RA, So HC, Leung GM, Peiris JM, Cowling BJ, Cauchemez S. Indirect protection from vaccinating children against influenza in households. Nature Communications. 2019;10(1):1–7.

16. Ribeiro-Rodrigues R, Kim S, da Silva FD, Uzelac A, Collins L, Palaci M, Alland D, Dietze R, Ellner JJ, Jones-López E, Salgame P. Discordance of tuberculin skin test and interferon gamma release assay in recently exposed household contacts of pulmonary TB cases in Brazil. PloS One. 2014:9(5).

17. Hill PC, Ota MO. Tuberculosis case-contact research in endemic tropical settings: design, conduct, and relevance to other infectious diseases. The Lancet Infectious Diseases. 2010;10(10):723–732.

18. Verver S, Warren RM, Munch Z, Richardson M, van der Spuy GD, Borgdorff MW, Behr MA, Beyers N, van Helden PD. Proportion of tuberculosis transmission that takes place in households in a high-incidence area. Lancet. 2004;363(9404):212–214.

19. Bennett DE, Onorato IM, Ellis BA, Crawford JT, Schable B, Byers R, Kammerer JS, Braden CR. DNA fingerprinting of Mycobacterium tuberculosis isolates from epidemiologically linked case pairs. Emerging Infectious Diseases. 2008;8(11):1124.

20. Brooks-Pollock E, Becerra MC, Goldstein E, Cohen T, Murray MB. Epidemiologic inference from the distribution of tuberculosis cases in households in Lima, Peru. Journal of Infectious Diseases. 2011;203(11):1582–1589.

21. Cohen T, Murray M, Abubakar I, Zhang Z, Sloutsky A, Arteaga F, Chalco K, Franke MF, Becerra MC. Multiple introductions of multidrug-resistant tuberculosis into households, Lima, Peru. Emerging Infectious Diseases. 2011;17(6):969–975.

22. Glynn JR, Guerra-Assunção JA, Houben RM, Sichali L, Mzembe T, Mwaungulu LK, Mwaungulu JN, McNerney R, Khan P, Parkhill J, Crampin AC. Whole genome sequencing shows a low proportion of tuberculosis disease is attributable to known close contacts in rural Malawi. PloS One. 2015;10(7) e0132840.

23. Yuen CM, Amanullah F, Dharmadhikari A, Nardell EA, Seddon JA, Vasilyeva I, Zhao Y, Keshavjee S, Becerra MC. Turning off the tap: stopping tuberculosis transmission through active case-finding and prompt effective treatment. The Lancet. 2015;386(10010):2334–2343.

